# Dissemination of dementia supporters and residents’ attitudes and recognition related to dementia in Japan: a municipal-level ecological study

**DOI:** 10.64898/2026.05.17.26353355

**Authors:** Taiji Noguchi, Kazushige Ide, Satoko Fujihara, Ayumi Kawagome, Masashige Saito, Katsunori Kondo, Toshiyuki Ojima

**Author notes:** Corresponding author: Taiji Noguchi, PhD, Department of Community Health and Preventive Medicine, Hamamatsu University School of Medicine, 1-20-1 Handayama, Chuoku, Hamamatsu, Shizuoka 431-3192.

## Abstract

**Background:** The Dementia Supporter Initiative is a national public education program in Japan that aims to foster positive attitudes and appropriate understanding of dementia to support people with Alzheimer’s disease and related dementia in the community. However, its influence on the community as a whole remains unclear.

**Objective:** This study examined the relationship between dementia supporter training and residents’ attitudes and recognition related to dementia at the municipal level.

**Methods:** This ecological cross-sectional study linked municipal-level data from the Japan Gerontological Evaluation Study 2022 wave with publicly available information on the number of dementia supporters. Residents’ beliefs and attitudes toward dementia and recognition of dementia consultation services were assessed by mail questionnaires and aggregated at municipal level. The proportion of dementia supporters in each municipality was calculated as of September 2022.

**Results:** Data from 69 municipalities were analyzed. The mean proportion of dementia supporters was 13.47% (2.62–44.85). A higher proportion of dementia supporters was positively correlated with community support-seeking for a family member with dementia (*r* = 0.328) and recognition of dementia consultation services (*r* = 0.501). Regression analysis adjusted for municipal covariates also showed their positive associations (per 10-percentage-point increase: coef. = 1.44, *p* = 0.047; coef. = 3.12, *p* < 0.001, respectively). No associations were observed with residents’ positive attitudes and appropriate understandings of dementia.

**Conclusions:** Wider dissemination of dementia supporters may contribute to better recognition of community support resources, but may be insufficient to influence broader public attitudes and understanding of dementia at the community level.

## Introduction

Dementia is a degenerative neurological disease that globally affects an estimated 54.7 million people in 2019, with a projected 152.8 million by 2050.^1^ The growing number of people living with dementia has substantial implications for individuals, their families, and healthcare systems.^2^ In response to these public health challenges, dementia-related policy has increasingly emphasized the importance of dementia-friendly cities and communities that enable people to live well with dementia by promoting supportive and inclusive communities.^3–6^ In Japan, a rapidly aging society, in which one third of the population is expected to develop dementia by 2060,^7^ the Basic Act on Dementia to Promote an Inclusive Society (Basic Act on Dementia) was enacted in 2023 to comprehensively enable people living with dementia to live with dignity and lead meaningful lives.^8^ A key to this legislation is to develop a society by deepening public knowledge and understanding of dementia for removing societal barriers to the social life of people living with dementia.

The dementia supporter (*Ninchisho Supporter*) is one of the practical policies to establish a dementia-friendly society.^9–12^ In 2005, the Japanese launched the “Japan National Campaign of Dementia Supporter Caravan.”^11–13^ This initiative, a representative measure to promote public understanding of dementia in Japan, aims to increase the number of residents who are capable of supporting people living with dementia and their families in the community, with efforts based on municipalities nationwide. Dementia supporters are not specialized professionals, but rather community members who provide kind support, such as watching over and encouraging people living with dementia, based on appropriate knowledge and understanding of dementia. By 2026, over 17 million people, ranging from children to older adults, have been trained as dementia supporters in Japan.^14^ This is one of the world’s largest initiatives to promote dementia-friendly practices in terms of population sizes.^10^ Inspired by this initiative in Japan and a similar model in the United Kingdom,^15^ almost 19 million people in 56 countries are currently trained and participating as “dementia friends,” including dementia supporters.^4^

The training of dementia supporters in Japan is mainly conducted on “dementia supporter training programs” of approximately 90 minutes. In this program, participants, including community residents, corporations, retailers, medical and care staffs, and schools, receive comprehensive education on basic knowledge of dementia, ways to care for people living with dementia, family support, and community consultation services provided by local governments.^13, 16^ This education program is delivered continuously by volunteers, “Caravan Mates,” who organize training lecture and serve as instructors, as they are trained and registered by the local governments. The contents of this program were updated in 2023 in response to the Basic Act on Dementia, shifting from a focus on medical knowledge alone to including the perspectives of living with the person concerned and encouraging participants to think of dementia as their own issue.^17^ Additionally, as a whole, the words and expressions were reconsidered to avoid fear and prejudice.^17^

Despite the international spread of dementia supporters, empirical evidence on the benefits of their training remains limited. Previous studies have reported improvements in positive attitudes and knowledge about dementia before and after the program among students^16, 18, 19^ and community residents,^20^ as well as correlations between program participation and attitudes and beliefs in cross-sectional comparisons with non-participants.^21,22^ However, these studies have mainly focused on short-term individual-level outcomes or cross-sectional associations at the participant level. In contrast, little is known about whether the dissemination of dementia supporters is reflected in dementia-related attitudes and recognition at the community level. From the perspective of dementia-friendly societies, it is crucial to examine collective outcomes, such as societal acceptance, support-seeking, and consultation behavior at the community level as well as the individual level.^3^ Furthermore, although the number of dementia supporters has increased substantially in Japan, the extent of dissemination may differ across communities. It is important to explore how such differences relate to residents’ beliefs and attitudes toward dementia, which can contribute to the evaluation and improvement of policies. These evaluations are particularly needed at the municipal level, the unit of implementation for dementia-related measures.^23^ However, it remains unclear how the status of dementia supporter training is related to residents’ attitudes and recognition of dementia at the municipal level.

Accordingly, this study aimed to examine the relationships between the status of dementia supporter training and residents’ attitudes and recognition related to dementia at the municipal level using an ecological analysis design. Specifically, we evaluated whether the municipal-level dissemination of dementia supporters was associated with residents’ positive beliefs and attitudes and appropriate understandings of dementia and recognition of the consultation services. This study provides a preliminary contribution as a policy evaluation of dementia supporter training in relation to dementia-friendly societies.

## Methods

### Study design and participants

This was a cross-sectional ecological study using data from the Japan Gerontological Evaluation Study (JAGES), a population-based survey of individuals aged 65 years and older in Japan,^24^ with municipal-level linkage of publicly available data on the status of dementia supporter training.^14^

Figure 1 shows the sample selection flow. Between November and December 2022, JAGES distributed self-administered questionnaires by mail or in person to older residents who were not certified as needing long-term care in 75 municipalities across 23 prefectures. These municipalities were not randomly selected, but they covered a wide range of regions and population sizes in Japan. Random sampling was used in 21 large municipalities, while all eligible residents were surveyed in 54 small municipalities. Among the 338,742 residents invited to participate, 228,119 returned the questionnaires (response = 67.3%). Of them, we excluded respondents whose age and/or sex could not be confirmed (n = 121), those certified as needing long-term care under the national systems or preventive long-term care services provided by municipality (n = 10,766), those for whom local government research cooperation was not available (n = 18,642), those who did not consent to study participation (n = 6,094), and those who were oversampled in one municipality (n = 388). Thus, data from 192,108 individuals across 73 municipalities were used (valid response = 56.7%).

**Figure 1.**
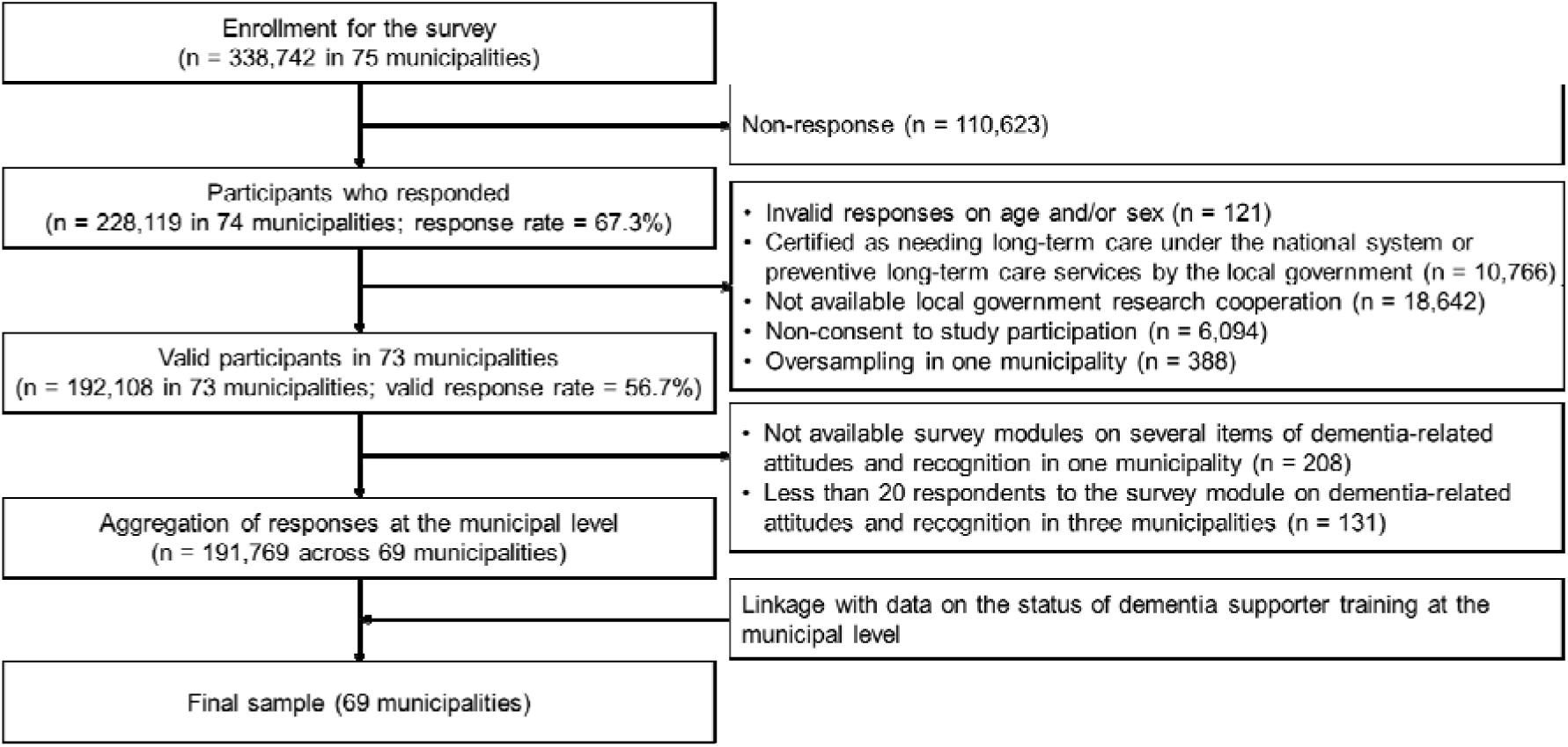
Sample selection flow.

Next, an aggregate dataset of responses on attitudes and recognition toward dementia at the municipality level was generated and linked to publicly available data on the status of dementia supporter training in municipalities.^14^ In this process, we excluded data from one municipality whose survey module on several items of dementia-related attitudes and recognition was unavailable (n = 208). Furthermore, to avoid the inaccuracy of the municipal-level data given the small sample size, we excluded three municipalities with fewer than 20 respondents (n = 131), because the relevant module had been administered to only approximately one-eighth of participants. Consequently, data from 69 municipalities comprising 191,769 respondents, with links to data on the status of dementia supporter training, were included in the analysis (mean number of observations per municipality = 2,779; range = 216 to 17,546).

This study was reviewed and approved by the ethics committees on human subjects at Chiba University (No. M10460) and Hamamatsu University School of Medicine (No. 91123). The mailed questionnaire was accompanied by an explanation of the study. Participants who agreed to participate were asked to check the consent box on the questionnaire and return the completed form; those who did so were considered to have provided informed consent. All our procedures were conducted in accordance with the Declaration of Helsinki.

### The status of dementia supporter training

To assess the dissemination status of dementia supporters, we used publicly available data on the implementation status of the Dementia Supporter Caravan provided by the Community-based Co-Operation Policy-Alliance of Local Governments.^14^ In line with the timing of the survey on residents’ attitudes and recognition related to dementia, we referred to the report as of September 2022 and extracted the total number of dementia supporters, including Caravan Mates, in each municipality. Based on the total population in municipalities from the Basic Resident Register (as of January 1, 2022), we finally calculated the proportion of dementia supporters per the total population (%), which was used as an indicator of the dissemination status in each municipality.

### Beliefs and attitudes toward dementia

Based on previous reports,^21, 25^ participants completed five items related to their beliefs and attitudes toward dementia: (i) preference for aging in place with dementia (“Do you agree that you would want to continue living in your own home with support from people around you if you developed dementia?”); (ii) beliefs about social engagement of people living with dementia (“Do you agree that people with dementia should have roles and participate in community activities?”); (iii) understanding of dementia-related symptoms (“Do you agree that behaviors such as shouting, aggression, or wandering in people living with dementia often occur when their needs are not being met?”); (iv) respect for autonomy of people living with dementia (“Do you agree that people living with dementia should be able to make their own everyday life decisions as much as possible?”); and (v) community support-seeking for a family member with dementia (“Do you agree that you would want neighbors and acquaintances to be aware so that they could provide support if a family member developed dementia?”). All items used a 5-point Likert scale ranging from “strongly disagree” to “strongly agree,” with a midpoint of “neither agree nor disagree.” For each item, the percentage of positive endorsement (“strongly agree” or “agree”) was aggregated at the municipal level (range: 0 to 100).

### Recognition of dementia consultation services

Participants answered the question, “Are you aware of any consultation services for dementia?” (response options: yes or no).^26^ The percentage of “yes” responses was aggregated at the municipality level (range: 0 to 100).

### Covariates

Population density, aging proportion (the proportion of people aged 65 and older), and fiscal strength index (calculated as the three-year average of the ratio of standard financial revenue to standard financial need) were used as municipal-level covariates. These data were obtained from the Japanese Census in 2022.^27^

### Statistical analysis

First, descriptive statistics of municipal-level residents’ attitudes and recognition related to dementia were calculated. Second, ecological correlations of the proportion of dementia supporters with residents’ attitudes and recognition of dementia in municipalities were analyzed. Third, to examine their relationships, multivariable linear regression analyses adjusted for municipal population density, aging proportion, and fiscal strength index were performed, and regression coefficients (coef.) and 95% confidence intervals (CIs) for each outcome were estimated. In these models, the proportion of dementia supporters was entered per 10-percentage-point increase. Because population density was highly right-skewed, a logarithmic transformation was applied before it was entered into the analytical model. Estimation was performed using robust standard errors to address potential heteroscedasticity across municipalities.

The significance level was set at < 0.05. All statistical analyses were conducted using R software (Version 4.5.2 for Windows; R Foundation for Statistical Computing, Vienna, Austria).

## Results

Data from 69 municipalities were analyzed. Table 1 shows the municipality characteristics. The mean proportion of dementia supporters in the population was 13.47% (standard deviation = 8.30). This ranged from 2.62% to 44.85%, representing a difference of approximately 42 percentage points. Regarding beliefs and attitudes toward dementia, around half of the municipalities showed positive endorsement for each item, except for the item of community support-seeking for a family member with dementia (mean proportion: 43.00 to 56.03). Meanwhile, around 70% of municipalities showed positive endorsement for community support-seeking for a family member with dementia. Across these items, the difference between municipalities was approximately 30 percentage points. Regarding recognition of dementia consultation services, the mean level was 35.26%, with a difference of approximately 30 percentage points between municipalities.

**Table 1.** The characteristics of the study municipalities (n = 69)

|  | Mean | SD | Median | Min | Max |
| --- | --- | --- | --- | --- | --- |
| Proportion of dementia supporters per population (%) | 13.47 | 8.30 | 10.72 | 2.62 | 44.85 |
| Population density (people/km<sup>2</sup>) | 1655.66 | 2476.16 | 599.17 | 1.18 | 10873.77 |
| Aging proportion (%) | 32.97 | 6.73 | 32.40 | 21.50 | 48.00 |
| Financial strength index | 0.63 | 0.32 | 0.60 | 0.16 | 1.81 |
| Beliefs and attitudes toward dementia (0-100) |  |  |  |  |  |
| Preference for aging in place with dementia | 56.03 | 5.32 | 55.80 | 40.00 | 75.00 |
| Beliefs about social engagement of people with dementia | 48.91 | 5.19 | 48.60 | 37.50 | 66.70 |
| Understanding of dementia-related symptoms | 54.77 | 5.05 | 55.00 | 42.20 | 74.10 |
| Respect for the autonomy of people with dementia | 43.00 | 4.94 | 42.90 | 30.90 | 66.70 |
| Community support-seeking for a family member with dementia | 72.24 | 5.74 | 71.90 | 60.30 | 83.70 |
| Recognition of dementia consultation services (0-100) | 35.26 | 5.93 | 35.30 | 24.40 | 54.20 |

SD, standard deviation.

Figure 2 presents the scatter plots and correlation analyses of the status of dementia supporter training and residents’ attitudes and recognition related to dementia at the municipal level. The proportion of dementia supporters showed little correlations with preference for aging in place with dementia (*r* = -0.001, *p* = 0.992), beliefs about social engagement of people with dementia (*r* = 0.075, *p* = 0.542), understanding of dementia-related symptoms (*r* = -0.014, *p* = 0.908), and respect for the autonomy of people with dementia (*r* = -0.021, *p* = 0.867). In contrast, the proportion of dementia supporters was positively correlated with community support-seeking for a family member with dementia (*r* = 0.328, *p* = 0.006) and recognition of dementia consultation services (*r* = 0.501, *p* < 0.001).

**Figure 2.**
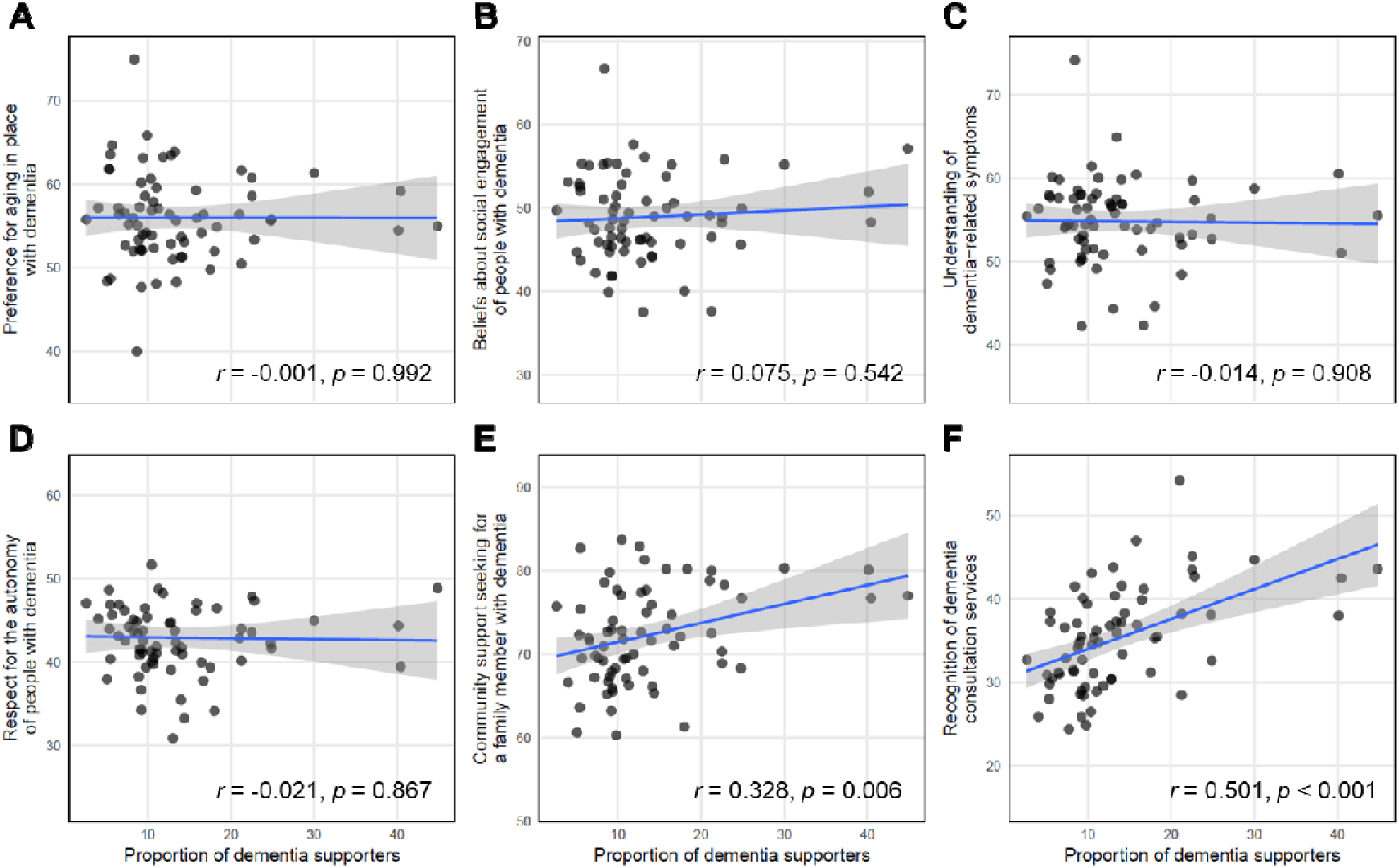
Correlations between the status of dementia supporter training and residents’ dementia-related attitudes and recognition. Scatter plots show the correlations between the proportion of dementia supporters and residents’ dementia-related outcomes at municipal level. Each dot represents an observation (municipality). Blue lines indicate fitted regression lines, and gray shaded areas indicate 95% confidence intervals. The outcomes shown are (A) preference for aging in place with dementia, (B) beliefs about social engagement among people with dementia, (C) understanding of dementia-related symptoms, (D) respect for the autonomy of people with dementia, (E) community support-seeking for a family member with dementia, and (F) recognition of dementia consultation services. Pearson’s correlation coefficients are shown.

Table 2 shows the association of the status of dementia supporter training with residents’ attitudes toward and recognition of dementia, based on linear regression analysis. After adjustment for municipal covariates, the proportion of dementia supporters was associated with greater community support-seeking for a family member with dementia (coef. = 1.44, 95% CI = 0.05 to 2.83, *p* = 0.047). Additionally, the proportion of dementia supporters was associated with greater recognition of dementia consultation services (coef. = 3.12, 95% CI = 1.47 to 4.77, *p* < 0.001). However, no associations were observed for the other dementia-related belief and attitude outcomes.

**Table 2.** Relationships between dementia supporter training and residents’ dementia-related attitudes and recognition, based on linear regression analysis.

|  | Crude model |  | Adjusted model |  |
| --- | --- | --- | --- | --- |
|  | Coef. (95% CI) | *p*-value | Coef. (95% CI) | *p*-value |
| Preference for aging in place with dementia | -0.01 (-1.17, 1.15) | 0.989 | 0.10 (-1.32, 1.52) | 0.889 |
| Beliefs about social engagement of people with dementia | 0.47 (-1.10, 2.04) | 0.562 | 0.51 (-1.21, 2.24) | 0.562 |
| Understanding of dementia-related symptoms | -0.09 (-1.39, 1.22) | 0.897 | -0.24 (-1.74, 1.25) | 0.752 |
| Respect for the autonomy of people with dementia | -0.12 (-1.56, 1.31) | 0.868 | -0.13 (-1.78, 1.51) | 0.874 |
| Community support-seeking for a family member with dementia | 2.27 (1.07, 3.48) | < 0.001 | 1.44 (0.05, 2.83) | 0.047 |
| Recognition of dementia consultation services | 3.59 (2.06, 5.11) | < 0.001 | 3.12 (1.47, 4.77) | < 0.001 |

Coef., unstandardized regression coefficient; CI, confidence interval.
The proportion of dementia supporters was scaled in 10-percentage-point units.
Estimates were obtained using robust standard errors and were adjusted for municipal population density (log scale), aging rate, and financial strength index.

## Discussion

This ecological study examined the relationship between the proportion of dementia supporters and residents’ attitudes and recognition related to dementia at the municipal level. Municipalities with a higher proportion of dementia supporters showed greater willingness to seek support from the community for a family member living with dementia and higher recognition of dementia consultation services. However, no associations were observed for residents’ values, beliefs, and understanding of dementia. These findings suggest that wider dissemination of dementia supporters may be related to more supportive community-level environments for people living with dementia and their families.

An important point is that the present study did not examine whether individuals who completed the dementia supporter training program themselves had more positive and favorable attitudes and behaviors. Rather, this study examined whether municipalities with greater dissemination of dementia supporters showed different patterns among residents as a whole. From this perspective, the association with recognition of dementia consultation services may indicate that municipalities with spreading dementia supporters have community-level support environments in which information on dementia-related resources is more familiar to residents. Additionally, considering that local social resources and consultation services, including community comprehensive support centers, are introduced within the program,^28^ the cumulative implementation of the program may contribute to residents’ recognition of dementia-related support resources. Meanwhile, the association with willingness to seek support from neighbors and acquaintances may reflect a municipal context in which asking for help is more acceptable in community life. This means that fostering an appropriate understanding of dementia through the program may reduce fear of and stigma toward dementia,^29, 30^ thereby lessening reluctance to disclose that a family member has dementia to others. Although the magnitude of the association was modest (per 10-percentage-point increase in the proportion of dementia supporters, 1.4-percentage-point increase for community help-seeking and 3.1-percentage-point increase for consultation service recognition), the effect sizes appear to be reasonable in light of previous research at the municipal level.^31^ Nevertheless, as these represent early steps of access to support, the spread of dementia supporters may still be relevant from the perspective of developing more supportive community environments.

No associations were observed for positive beliefs and attitudes toward dementia or understanding of dementia. One possible explanation is that the dementia supporter training program is a relatively brief (approximately 90 minutes) and usually one-time education program, and such a format may be insufficient to substantially change residents’ values and beliefs at the community level. Previous studies suggest that contact and interaction with people living with dementia in addition to lectures, are associated with reduced stigma and improved knowledge related to dementia.^30, 32^ Yet, knowledge of the disease alone may actually increase fear, suggesting the importance of personhood-based knowledge.^33, 34^ To influence attitudes toward dementia broadly at the municipal level, person-centered programs that include interactions with people living with dementia and simulated experiences may be necessary. Whereas, in the present study, the average proportion of dementia supporters was approximately 13% of the population. Although there is no established threshold for sufficient dissemination, this level of penetration may not have been sufficient to influence the beliefs and attitudes of the whole community, given that many of the trainees have not subsequently engaged in related activities.^12^ Nevertheless, in 2023, the program in Japan was updated to include more of the perspectives of those involved and their families and to reduce misunderstanding and prejudice.^17^ Although the present study, which used data collected before this revision, could not evaluate its effects, this change may contribute to fostering more positive values and beliefs for the entire community in the future. Furthermore, the dissemination of dementia supporters varied substantially across municipalities, suggesting that the spread of the program remains uneven. These results indicate that both qualitative improvement and quantitative expansion of dementia supporters are required to achieve broader influences of residents’ values and perceptions toward dementia at the community level.

From a policy perspective, this study highlights the importance of evaluating and monitoring dementia-related measures. The Dementia Supporter Caravan is an initiative intended to promote positive attitudes and appropriate understanding of dementia among the public and is positioned as an implementation measure to foster a “new perspective on dementia” in line with the Basic Act on Dementia in Japan.^8, 35^ To achieve these objectives, it is necessary to verify the benefits of dementia-related measures and to improve them based on evidence, which requires assessment and monitoring of population attitudes, stigma, understanding, and knowledge related to dementia. Furthermore, considering that attitudes and stigma from society can adversely affect individuals living with dementia and their families, it is also essential to clarify how such measures may benefit those affected and their families.

There are several limitations in this study. First, the ecological study design could not address causal relationships at the individual level because of ecological fallacy. It should be noted that municipality-level associations do not necessarily reflect individual-level relationships. Further research, including multilevel analysis, is needed to examine the relationship between the dissemination of dementia supporters in municipalities and individual attitudes and recognition. Second, the number of dementia supporters per municipality, identified from publicly available data, reflects the number of program participants but may include repeated attendance by the same individual. Therefore, it may not necessarily correspond to the actual number of unique supporters in each municipality, although it may serve as an indicator of the degree of dissemination. Third, whereas the status of dementia supporter training was based on information from all generations, attitudes and recognition related to dementia in municipalities were evaluated using aggregated responses from older residents. This mismatch may have caused measurement error. Finally, the municipalities included in the analysis accounted for less than 5% of all municipalities in Japan, which may limit the generalizability of the findings. Still, recruitment was conducted nationwide and covered nearly half of the prefectures in Japan (23 out of 47).

## Conclusion

This study examined the ecological relationships between the proportion of dementia supporters and residents’ attitudes and recognition related to dementia at the municipal level. The dissemination of dementia supporters was positively associated with greater community support-seeking for family members living with dementia and with recognition of dementia consultation services in municipalities, but not with positive attitudes toward and understanding of dementia. These findings suggest that wider dissemination of dementia supporters may be related to more supportive community-level environments for people living with dementia and their families, warranting further investigation into the influence of dementia supporter training on residents’ attitudes and recognition of dementia.

## Acknowledgments

We wish to express our sincere gratitude to the staff in the surveyed municipalities for their contributions. We also thank the members of the 300 BM and AFC working group of the Japan Gerontological Evaluation Study.

## Author contributions

TN conceptualized and designed the study, analyzed the data, interpreted the results, and drafted and revised the manuscript. KI, SF, and AK helped to conceptualize and design the study, interpret the results, and reviewed and critically revised the manuscript. MS helped to conceptualize the study, participated in data collection and study design, supervised, and reviewed and critically revised the manuscript. KK, the principal investigator of the JAGES, helped to conceptualize to the study, participated in data collection and study design, supervised, and reviewed and critically revised the manuscript. TO helped to conceptualize the study, participated in data collection, supervised, and reviewed and critically revised the manuscript.

## Funding statement

This work was supported by the Japan Society for the Promotion of Science (JSPS), KAKENHI Grant Numbers (24K20158, 26H00568). The JAGES was supported by JSPS Grant-in-Aid for Scientific Research (20H00557, 20K10540, 21H03196, 21K17302, 22H00934, 22H03299, 22K04450, 22K13558, 22K17409, 23H00449, 23H03117, 23K21500), Health Labour Sciences Research Grants (19FA1012, 19FA2001, 21FA1012, 22FA2001, 22FA1010, 22FG2001), Research Institute of Science and Technology for Society (JPMJOP1831) from the Japan Science and Technology (JST), a grant from Japan Health Promotion & Fitness Foundation, TMDU priority research areas grant, and National Research Institute for Earth Science and Disaster Resilience. The views and opinions expressed in this article are those of the authors and do not necessarily reflect the official policy or position of the respective funding organizations.

## Statements and declarations

TN, KI, SF, AK, MS, KK, and TO have received compensation from the Community-based Co-Operation Policy-Alliance of Local Governments. The authors declare no other potential conflicts of interest with respect to the research, authorship, and/or publication of this article.

## Consent for publication

Not applicable.

## Data availability

For the JAGES, all inquiries are to be addressed to the data management committee via. All JAGES datasets have ethical or legal restrictions for public deposition because of the inclusion of sensitive information about human participants. The status of dementia supporter training is publicly available by the Community-based Co-Operation Policy-Alliance of Local Governments: https://www.caravanmate.com/

